# Temporal trends in public mass shootings: high-capacity magazines significantly increase fatality counts, and are becoming more prevalent

**DOI:** 10.1101/2019.12.12.19014738

**Authors:** Sherry Towers, Danielle Wallace, David Hemenway

## Abstract

**Background:** There has been disagreement in the literature regarding whether public mass shootings are becoming more frequent, or more deadly, and whether high-capacity magazine (HCM) weaponry is associated with increased fatalities.

**Objective:** We examined the 85 non-felony- and non-gang-related mass shootings with at least four people shot and killed that occurred in public places between 1995 to 2018 to examine the frequency and deadliness of public mass shootings, as well as the role of HCM weaponry in fatalities.

**Findings:** The per-capita rate of public mass shootings with at least four people killed has not significantly increased since 1995 (Negative Binomial linear regression p=0.20), but public mass shootings with at least six people killed have been increasing by 6.9% per year (p=0.01). Public mass shootings are becoming significantly more deadly, with fatalities per incident increasing by 4.5% per year (truncated Negative Binomial log-linear regression p<0.001).

Use of HCM weaponry banned during the 1994 to 2004 Federal Assault Weapons Ban (FAWB) increased fatality counts on average by 1.9 times, and total number shot by 3.3 times (non-parametric bootstrap comparison of means (NPBCM), p < 0.001 in both cases). In these incidents, HCM pistols were as deadly as assault rifles. The fraction of firearms involving HCMs used by perpetrators more than doubled after the lapse of the federal ban, from 25% during the ban to 59% after (Binomial factor regression, p<0.001).

Our online visual analytics application, https://mass-shooting-analytics.shinyapps.io/mass_shootings/, allows interested parties to examine and download our data, replicate our analysis results and explore the analysis with alternate selections.

**Policy recommendations:** A re-instatement of the federal ban on high-capacity magazines in pistols and rifles, or special taxes on such weaponry, may reduce public mass shooting fatalities.

## Introduction

High-fatality public mass shootings are a rare form of firearm violence, but occur more often in the US than in any other developed country (Lankford, 2016, 2019). Previous analyses have noted that mass shootings are more likely than other types of firearm violence to receive national media attention (Duwe, 2000; Silva & Capellan, 2019). Massacres that occur in public places, particularly those that are seemingly random events perpetrated by a stranger to the victims, tend to capture the attention of the public, often provoking feelings within the public audience of “that could have been me” (Petee, Padgett, & York, 1997; Wayment, 2004). Such events often trigger vigorous public debate about policies that address all of the problems of firearm violence (Barry, McGinty, Vernick, & Webster, 2013; Bowers, Holmes, & Rhom, 2010; Krouse & Richardson, 2015). Unfortunately, such debate is often based on partisan rhetoric instead of evidence-based research (Cook & Goss, 2014).

While much has been written on the subject of mass shootings [e.g. (Duwe, 2007; Fox, Levin, & Fridel, 2018; Klarevas, 2016)], there has been a remarkable dearth of peer-reviewed, non-partisan statistical analyses of the dynamics that underlie the frequency and casualty-counts of such events, perhaps due to what has amounted to an effective Congressional moratorium on funding to the Centers for Disease Control and Prevention (CDC) for studies of firearm violence, in place since 1996 (Kellermann & Rivara, 2013; Winker, Abbasi, & Rivara, 2016). The few analyses that have been performed often disagree on various aspects of mass shootings (Cohen, Azrael, & Miller, 2014; Klarevas, Conner, & Hemenway, 2019; Lankford & Tomek, 2018; Towers, Gomez-Lievano, Khan, Mubayi, & Castillo-Chavez, 2015; Towers, Mubayi, & Castillo-Chavez, 2018), including whether the frequency of such events has been increasing (Cohen et al., 2014; Lott, 2015), and/or whether certain types of weaponry, like assault rifles, play a significant role in casualty counts (Kleck, 2016; Lemieux, 2014). Poorly executed statistical analyses and non-transparency in data selection have appeared to play a role in some of the disagreements (Towers et al., 2018); indeed, a recent study by RAND researchers has noted a lack of statistical rigor in many analyses of firearm violence (Schell, Griffin, & Morral, 2018).

To examine the frequency and deadliness of public mass shootings, as well as the role of HCM weaponry in fatalities, we focused on mass shooting incidents with at least four people shot and killed (not including the perpetrator) that occurred in public places; victims shot and killed in private homes or on private land were not included in the tally of fatalities. Additionally, the Federal Assault Weapons Ban (FAWB) that was in effect from September 13, 1994 to September 13, 2004, and its subsequent lapse, might plausibly have had an effect on the types of weapons used in mass shootings (Koper & Roth, 2001; Koper, Woods, & Roth, 2004), which may also have impacted the number of fatalities per incident. We thus examined public mass shooting events from the beginning of the FAWB to the end of 2018, and examined whether the fraction of events that involved weaponry that had been banned during the FAWB differed significantly during versus after the ban and whether the number killed in each event was related to whether or not such weaponry was used.

## 2 Methods and Materials

### 2.1 Data

No complete and comprehensive official database of US public mass shootings currently exists. Previous researchers have often relied on incomplete government databases, or databases compiled by unofficial sources such as USA Today and Mother Jones (Fox & Fridel, 2016). Like other researchers, we have found various compilations of public mass shootings often to be incomplete or have errors in the event details, and/or have apparent inconsistencies in criteria for inclusion (Fox & Levin, 2015; Huff-Corzine et al., 2014; Klarevas, 2016; Towers et al., 2015). In our analysis we have attempted to rectify this problem by compiling a comprehensive list of all high-fatality public mass shootings in the United States from 1995 onward that satisfy a well-defined set of criteria.

To begin, we define “mass murder” according to the 2010 Federal Bureau of Investigation (FBI) criteria of four or more killed (Huff-Corzine et al., 2014), since it is these high-fatality count events which are most likely to receive widespread media attention, and foment debate on potential changes to state and federal policies to address the overall problem of firearm violence in the US (Duwe, 2000; Silva & Capellan, 2019).

Events included in our analysis may have had multiple perpetrators, and may have occurred in multiple public locations, but must have occurred within a 24-hour period to be included. Shootings related to gang violence or other criminal activity were excluded, as were shootings that could plausibly have been incited for reasons of self-defense, and shootings that began with a police encounter during a traffic stop precipitated by suspicion of illegal activity. Only events with at least four people shot and killed in public places (the four fatalities did not include the perpetrator) were included; victims killed during the incident on private property were not included in the fatality counts. We also only counted as fatalities victims who died within one month of the incident, because it is the initial fatality count that primarily captures the public attention.

As detailed in Appendix A, sources of information used in this analysis included the FBI Supplementary Homicide Reports, along with many past print and online compilations of mass shootings collected by researchers and various public and private organizations. From these sources of data, events were selected that fit our definition of high-fatality public mass shootings. While there was significant overlap in many of the past compilations of mass shootings, events were often included in some compilations that were not included in others, in part due to differing inclusion criteria in the compilations (Klarevas, 2016). However, we also found several instances where events should have likely been included in some compilations (according to the stated event inclusion criteria) but were excluded without explanation as to why, and it is unclear whether the exclusion was deliberate, or instead due to ignorance of the incident on the part of the researchers compiling the list.

To avoid this confusion with our database, we created a secondary database of events that have been listed as “public mass shootings” in some past compilations, but did not meet the criteria for inclusion in our primary database (because, for example, all or most of the people killed were killed in private homes rather than public places, or only some of the victims were killed by firearms). An example of an event typically tallied as a “public mass shooting” by several past compilations, but was excluded in ours, is the September 2, 2008 incident in Alger, WA, where the perpetrator killed five people in private homes, then shot and killed one other person during a subsequent public shooting spree. We included such events in our secondary database, with explanation as to the specific criteria each event did not meet for inclusion in the primary data. In our online visual analytics application, available at https://mass-shooting-analytics.shinyapps.io/mass_shootings/, we allow the user to repeat our analyses with the option to specifically include or exclude certain events. The visual analytics application also contains an extensive set of links to online material that were used as sources of information related to each event.

We selected incidents from the beginning of the FAWB on September 13, 1994 to the end of December 2018. However, because no incidents meeting our criteria occurred from September 13, 1994 to the end of 1994, the data selection effectively is for events between 1995 to 2018. We found 85 public mass shootings.

For each incident, we examined media reports, court documents, and other documentation to determine the number and types of weapons used, including whether weaponry banned during the FAWB was among the weaponry used, such as high-capacity magazines carrying more than ten rounds of ammunition (used in either a rifle or pistol), or an “assault rifle”. We use the definition of assault rifle specified in the FAWB legislation, which are semi-automatic rifles with a detachable magazine that possess two or more of the following features; folding or telescoping stock, pistol grip, bayonet mount, flash suppressor or threaded barrel, or grenade launcher (see https://bit.ly/2U6dvOS, accessed November, 2019). For 77 (90.6%) incidents we were able to determine whether or not such weaponry was used.

### 2.2 Statistical methods

#### 2.2.1 Temporal trends in frequency of incidents

Mass shooting events, per-capita, are rare, and thus Ordinary or Generalized Least Squares methods are inappropriate for analyses of the trends in the frequency of such events (Gardner, Mulvey, & Shaw, 1995). Also, when analyzing the temporal trends in the frequency of mass shootings over a long time period, the effect of population change must be taken into account (for example, everything else being equal, doubling the population size will double the frequency of mass shootings). In addition, binning of data into a relatively long time period, like a year, necessitates loss of information and reduces sensitivity of the analysis (Towers et al., 2018). In this analysis, we thus performed an un-binned likelihood fit to the data (beyond the necessary binning of data into integer days), and examined the temporal change in the per-capita number of mass shootings per day, *λ/N*, where *N* is the population size, and *λ* is the average number per day. We employed a population-standardized log-linear model (Osgood, 2000), with the predicted number per day, λ, described as log λ = log *N* + *β*_0_ + *β*_1_*t*, where *t* is the date. To account for potential over-dispersion in the count data involved in our analyses, we used a Negative Binomial likelihood fit (Lloyd-Smith, 2007).

#### 2.2.2 Temporal trends in fatalities per incident

The number of fatalities per incident is integer count data, and Least Squares methods are again not appropriate for statistical analyses of such data because they are often highly non-Normal and skewed (Osgood, 2000). While some previous analyses have used Poisson regression methods to analyze mass shooting fatalities (e.g. Gius, 2015, 2018), this does not take into account the gross over-dispersion in the distribution of fatalities, and can thus lead to over-estimation of the statistical power of the analysis. Negative Binomial likelihood methods are more appropriate for over-dispersed count data (Lloyd-Smith, 2007), but in an analysis like ours the model must also account for the fact that the data are truncated by the selection of the minimum number killed in order to enter the sample (in the case of this analysis, four). As described in Appendix A in the supplementary material, we employed a Negative Binomial likelihood fit with the number killed, k, left-truncated at k ≥ 4.

#### 2.2.3 Comparison of the number of fatalities per incident for events that do, and do not, involve weaponry banned during the FAWB

Because mass shootings are rare, and the number of fatalities per incident are integer data truncated at k ≥ 4 fatalities, hypothesis tests based on assumptions of Normality, such as the Student’s t-test are not appropriate for comparing the means of the distributions (Nair, 1941). The sample sizes are also generally not large enough such that the Central Limit Theorem applies, such that the means can be compared using the Z test. In addition, non-parametric tests that compare distributions, such as the Kolmogorov-Smirnov test, are not appropriate in the presence of ties (i.e., many overlapping identical values in the data sets) (Corder & Foreman, 2014).

Here we relied on non-parametric bootstrapping methods to compare the means of distributions of fatalities (Barber & Thompson, 2000; Mooney, Duval, & Duvall, 1993). For events that do, and do not, involve weaponry banned during the FAWB, we drew 10,000 bootstrapped distributions, sampled with replacement from the original distributions. For each sampled distribution we calculated the mean, and determined how often the mean of the distribution of events that did not involve such weaponry was greater than the mean of the distribution of events that did. This fraction forms the p-value for the one-sided comparison test (Barber & Thompson, 2000; Mooney et al., 1993).

## 3 Results

### 3.1 Temporal trends in frequency of incidents

In Figure 1 we show the temporal trends in the per-capita rate of all public mass shootings for selections of at least four, six, and eight killed, respectively. For the sample with ≥4 killed, there is no significant linear trend in per-capita rate, but both the ≥6 and ≥8 killed samples show significant increases in time in the per-capita rates of 6.9% and 13.6% per year, respectively (population standardized Negative Binomial log-linear regression p-values p=0.01 and p=0.001, respectively).

**Figure 1:**
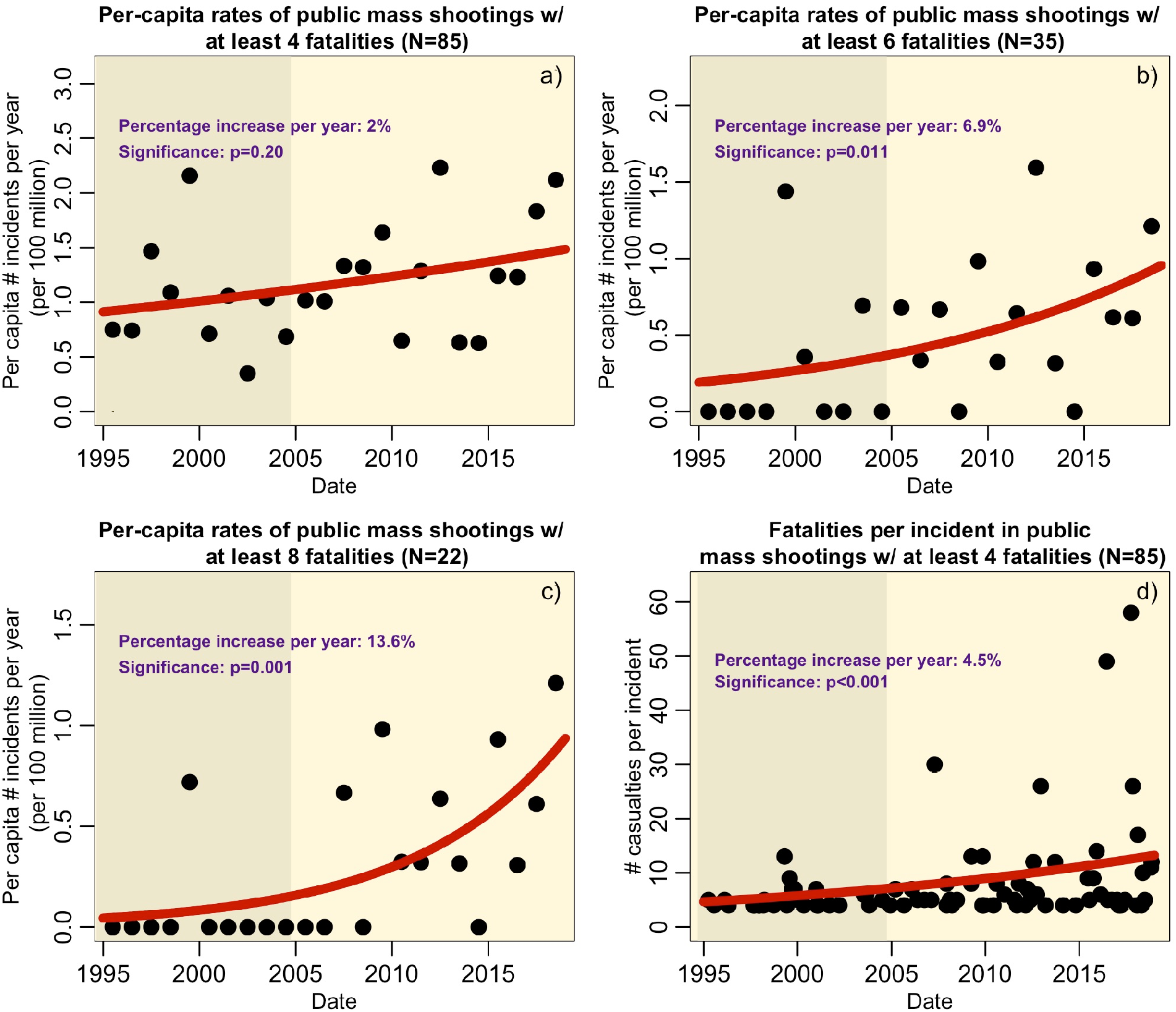
Plots a), b) and c) show the temporal trends in the per capita rate of public mass shootings, for selections of at least four, six, and eight people killed, respectively (note: for display purposes the data are binned by year, but the fit is to the number of incidents per day). Overlaid in red are the results of population standardized Negative Binomial log-linear un-binned likelihood fits. The grey shaded region indicates the period of the Federal Assault Weapons Ban. Plot d) shows the temporal trends in the number of casualties per incident.

### 3.2 Temporal trends in fatalities per incident

During the FAWB there was an average of 5.2 fatalities per incident, compared to 9.3 after the lapse of the FAWB (1.8 times higher, non-parametric bootstrap p<0.001) (Figure 1). There has been a significant relative increase in the number of fatalities per incident of 4.5% per year since 1995 (truncated Negative Binomial log-linear regression p-value p < 0.001).

There is no significant temporal trend in the number of fatalities in incidents that do not involve HCM weaponry (truncated Negative Binomial log-linear regression p-value p=0.11).

### 3.3 Effect of weaponry types on the number of fatalities per incident

For the 77 out of 85 events for which the type of weaponry was known, the average number of fatalities in the 23 events that did not involve HCM weaponry banned during the FAWB (including high-capacity semiautomatic pistols) was 5.0. The 54 events that involved HCM weaponry had on average 9.8 fatalities (1.9 times higher, non-parametric bootstrap comparison of means p < 0.001) (Figure 2).

**Figure 2.**
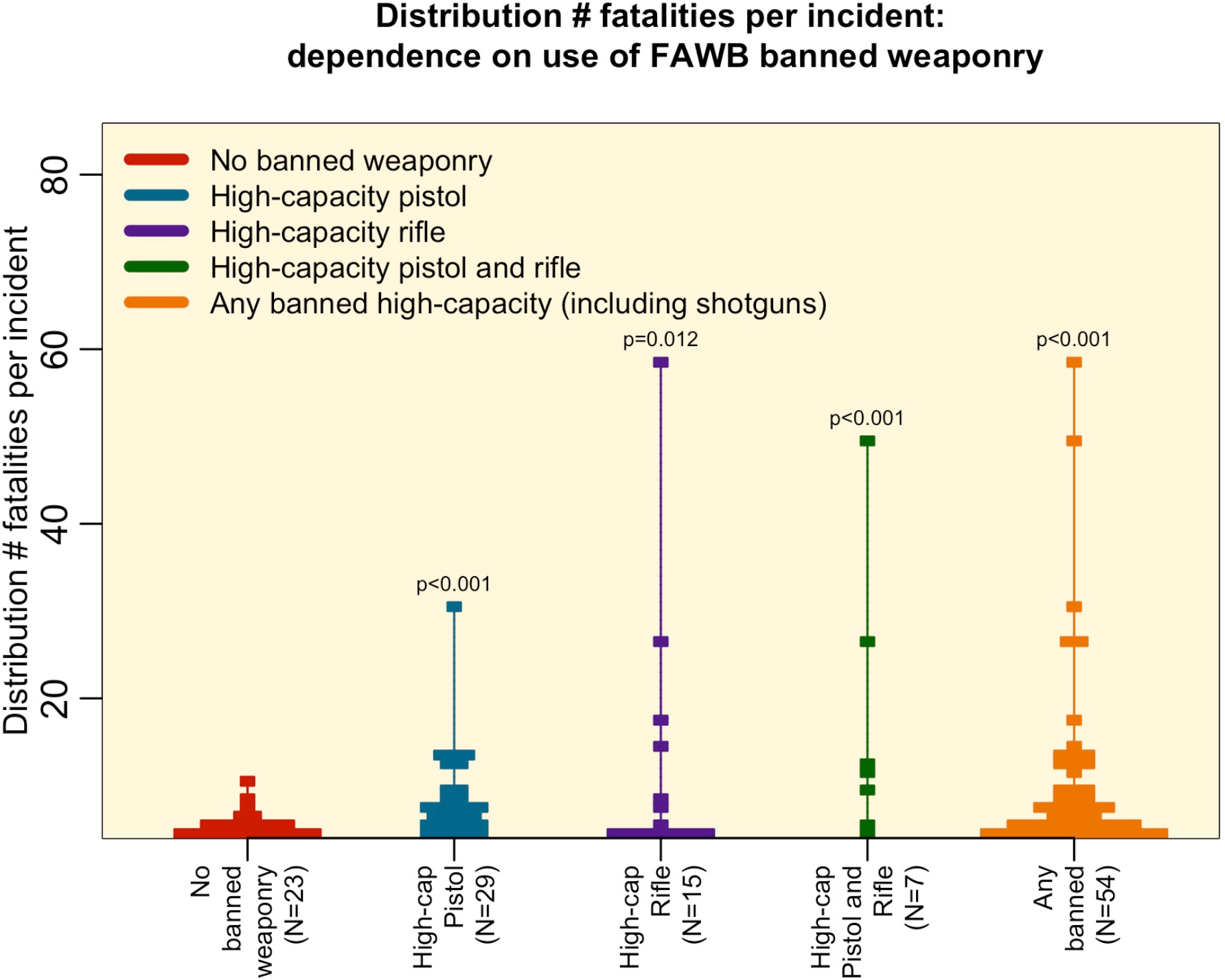
Distribution of the number of fatalities in public mass shootings between 1995 to 2018 with ≥4 people killed, by type of weaponry used. The width of the bars is proportional to the number of events in each group with a particular number of fatalities. The p-values represent the one-sided comparison tests of the means of the distribution of fatalities for events that involved banned weaponry to the mean of distribution of the events that did not involve such weaponry.

The difference in the means is not solely due to ultra-high fatality count events such as the October 1, 2017 Las Vegas, NV massacre (58 fatalities) and the June 12, 2016 Orlando, FL Pulse nightclub shooting (49 fatalities); excluding those two events from the incidents involving HCM weaponry yielded an average of 8.1 fatalities (1.6 times higher, non-parametric bootstrap comparison of means p < 0.001).

The difference in the total number shot (some of whom survived) is also significant. The average number of people shot in the 23 events that did not involve HCM weaponry was 7.5, compared to an average of 25.0 shot in events that involved HCM weaponry (3.3 times higher, p < 0.001). Excluding the October 1, 2017 Las Vegas, NV massacre and the June 12, 2016 Orlando FL Pulse nightclub shootings yields an average of 14.8 people shot (2.0 times higher compared to incidents perpetrated without HCM weaponry, p < 0.001).

Even just the use of high-capacity magazine pistols (and not an assault rifle) resulted in significantly more fatalities on average compared to that involved no banned weaponry (1.6 times higher, non-parametric bootstrap comparison of means p<0.001) (Figure 2). Indeed, there was no significant difference in the number of fatalities between events for which the banned weaponry only involved an assault <u>rifle</u> compared to events for which the banned weaponry only involved a high-capacity semi-automatic pistol; 11.6 versus 8.0 killed on average, respectively (non-parametric bootstrap comparison of means p=0.79).

The number of fatal victims was significantly associated with the number of HCM firearms used in the incident (Negative Binomial linear regression p<0.001) [and was almost significant with the number of non-HCM firearms (p=0.06)]. This conclusion held even when the October 1, 2017 Las Vegas NV massacre was excluded. For incidents where only one firearm was involved, the average number of fatalities in incidents with a non-HCM firearm was 4.6, compared to 7.3 when an HCM firearm was used (non-parametric bootstrap comparison of means p<0.001).

### 3.4 Trends in the use of weaponry banned during the FAWB

During the FAWB, 48% of the public mass shooting events involved weaponry banned during the FAWB. After the lapse of the ban, 71% of such events involved weaponry that would have been banned during the FAWB (p=0.05, Binomial factor regression).

The average number of weapons per incident was not significantly different during and after the ban (2.2 versus 2.3 weapons per incident, non-parametric bootstrap comparison of means p=0.46). However, the average number of weapons involving HCMs per incident rose over two-fold, from an average of 0.6 during the ban, to 1.4 after the ban (non-parametric bootstrap comparison of means p=0.002). The fraction of firearms involving HCMs used by perpetrators also more than doubled after the lapse of the ban, from 25% during the ban to 59% after (Binomial factor regression, p<0.001). The significant differences remained even when the October 1, 2017 Las Vegas, NV massacre was excluded from the data set.

The differences during- and post-ban appear to have been partly driven by a rising popularity in use of high-capacity semiautomatic pistols by perpetrators. The average number of assault rifles per incident was 0.2 during the ban, compared to 0.7 after (0.3, excluding the October 1, 2017 Las Vegas NV massacre), non-parametric bootstrap comparison of means p=0.09 (p=0.24). In contrast, the average number of high-capacity semi-automatic pistols per incident during the ban was 0.3, compared to 0.6 post-ban (p=0.009). During the FAWB 13% of firearms used where HCM pistols, compared to 28% after the lapse (p=0.031, Binomial factor regression).

## 4 Discussion

In this analysis we employed rigorous statistical methodologies and well-defined selection criteria to examine high-fatality public mass shootings in the US. We have made efforts to ensure full transparency in our data selection criteria, and to make our data, referenced sources of information, and analysis methodologies publicly available (see https://mass-shooting-analytics.shinyapps.io/mass_shootings/). Our online interface to the database of public mass shootings used in this analysis is extensively annotated with links to external sources of information we used to determine event details, including whether weaponry banned during the FAWB was used by the perpetrator(s).

In our data collection, we found that several sources of mass shooting data commonly used by other researchers appeared to have errors, omissions, and inconsistencies— highlighting the importance of cross-checking and validating information when using data compiled by other investigators—on any issue.

In our analysis we focused on events with four or more people killed, since high-fatality incidents are most likely to receive significant amounts of national media attention (Duwe, 2000; Silva & Capellan, 2019), and are most likely to foment changes to public policy (Fox & DeLateur, 2014; Schildkraut & Elsass, 2016). Selecting events based on an arbitrary number of fatalities has been criticized, since people who were wounded in an attack were clearly meant to be killed by the perpetrator but instead managed to survive the attack (Capellan & Gomez, 2018; Lott & Landes, 2000). In addition, “thwarted” attacks, where the perpetrator apparently meant to shoot many people but was prevented from doing so by some means (such as bystander interference, for example) may provide information on whether or not various potential intervention measures are actually efficacious. While it might be well motivated to examine all shootings or attempted shootings, there may be biases in data selection and a lack of details related to the events. Even the FBI Supplementary Homicide Reports are incomplete in their details and in their tally of multiple-victim shootings (Huff-Corzine et al., 2014; Towers et al., 2015). Researchers currently must rely on media reports and online documents to attempt to fill in the gaps of the official record (Capellan & Gomez, 2018; Parkin & Gruenewald, 2017), and under-reporting by the media of low-fatality events can bias the detection efficiency for such events (Duwe, Kovandzic, & Moody, 2002; Schildkraut, Elsass, & Meredith, 2018; Silva & Capellan, 2019). In addition, the further back in time one goes, the more likely low-fatality events will escape notice.

Indeed, in our analysis, after an exhaustive search of public mass shootings from 1995 to 2018, we found events in our data set with four people killed that had not been included in tallies compiled by other researchers, indicating that lower-fatality incidents are likely easily over-looked in data collection efforts.

Various definitions of ‘high-fatality’ mass shootings have been used in the literature, with some researchers favoring a cutoff of even higher than four killed [e.g., (Klarevas, 2016; Kleck, 2016)]. For transparency, our online visual analytics application allows everyone to explore the trends in our data with more restrictive selections on the minimum number killed, and/or excluding or including specific events (see https://mass-shooting-analytics.shinyapps.io/mass_shootings/).

### 4.1 Temporal trends in frequency of incidents

Using an un-binned population standardized Negative Binomial likelihood fit, log-linearly regressing on time, we found that the per-capita rate of all public mass shootings with at least four killed has not significantly increased since 1995 (2% average relative increase per year, p=0.20), but the rate with six or more killed has (6.9% average relative increase per year, p=0.011).

Our finding that per-capita rates of public mass shootings have not increased significantly is in conflict with the results of a report (Cohen et al., 2014) using data from Mother Jones. However, we find 64 public mass shootings satisfy our selection criteria from 1995 to the end of 2014, while Mother Jones counted only 50 incidents over the same time period. Our findings are also different than (Krouse & Richardson, 2015), who examined public mass shootings with four or more individuals killed between the years 1970 to 2014. However, that analysis did not assess per-capita annual rates because it did not take into account the change in the US population over that time period and did not test the null hypothesis of no temporal trend in per-capita rates.

By contrast, our findings that the per-capita rates of very high-fatality incidents are increasing is in qualitative agreement with the results of (Klarevas, 2016; Klarevas et al., 2019) that examined mass shootings (not just public mass shootings) with six or more killed, and found that in the ten year period after the lapse of the FAWB there was a 183% increase in the rate of such massacres.

### 4.2 Temporal trends in fatalities per incident

We found that the number of fatalities per incident has significantly increased by 4.5% per year (truncated Negative Binomial log-linear regression p-value p < 0.001), and that the number of fatalities per incident was 1.8 times higher after the lapse of the FAWB than it was during it (non-parametric bootstrap p<0.001). This is in qualitative agreement with the results of both (DiMaggio et al., 2019) and (Klarevas, 2016).

### 4.3 Effect of weaponry types on the number of fatalities per incident

We found that the use of weaponry that had been banned during the FAWB significantly increased fatality counts in public mass shootings; the average number of fatalities in the 31 events that did not involve such weaponry (or for which it was unknown whether such weaponry was involved) was 4.8 while the 54 events that involved weaponry banned during the FAWB (including high-capacity semiautomatic pistols) had on average 9.8 fatalities (2.0 times higher, non-parametric bootstrap comparison of means p<0.001). The total number shot was over 3 times higher in incidents that involved HCM weaponry (non-parametric bootstrap comparison of means p<0.001).

This finding is in disagreement with the arm-chair reasoning of (Kleck, 2016) and (Fox & Fridel, 2016) who pointed out that many mass shooters are armed with multiple weapons, so high-capacity magazines would thus not likely make a difference since the shooter would not have to stop to reload. We found that fatality counts were significantly associated with the number of HCM weapons used, but were not significantly associated with the number of non-HCM weapons used. We also found that in incidents where only one firearm was used, the use of a HCM firearm significantly increased fatality counts, by a factor of 1.6 times. Similiar to (Klarevas et al., 2019), our findings highlight the importance of high capacity magazines as a likely factor in increasing the number of deaths.

### 4.4 Trends in the use of weaponry banned during the FAWB

During the FAWB, 48% of all public mass shooting events involved HCM weaponry. After the lapse of the ban, 71% of events involved such weaponry (p=0.047, Binomial factor regression). These results are in contrast to the claim made by (Krouse & Richardson, 2015) that only 27% of assailants in public mass shootings between 1999 and 2013 used weaponry that had been banned during the FAWB. Because (Krouse & Richardson, 2015) did not provide incident-level details of the 66 events in their analysis. We thus could not determine where the differences lie between their analysis and ours. We counted 51 public mass shooting events over the same time period, and we were able to determine the types of weaponry used for 45 cases, 31 of which (69%) involved weaponry that had been banned during the FAWB.

Our results also lie in sharp contrast with the claim by (Kleck, 2016) that “less than one third of 1%” of assailants in mass shootings used HCMs—a claim that was not substantiated by any cited sources of data. Our results also contrast with the claim by (Duwe, 2007) that “only 2%” of mass murders are committed with an assault weapon. It is unclear how Duwe arrived at this number, what time period was examined, how “mass murder” was defined in the analysis, and how it was determined whether or not an assault weapon was used. Their definitions of mass shootings are clearly very different from the one used in this paper; this again emphasizes the need for researchers of mass killings to be more transparent in specifying their data selection criteria.

The temporal trends we noted in the types of weaponry used in public mass shootings mirror the trends noted by (Koper, Johnson, Nichols, Ayers, & Mullins, 2017) who found that since the lapse of the FAWB high-capacity pistols and rifles are increasingly used in mass murders and other types of crime.

Our finding that the increasing use of formerly banned weaponry is largely driven by a increase in the popularity of high-capacity semi-automatic pistols among perpetrators, rather than assault rifles, is in agreement with the findings of (Klarevas, 2016; Klarevas et al., 2019), who found that the fraction of mass shootings involving six or more victims committed with an assault rifle did not significantly change post-ban, but the use of pistols involving HCMs did.

## 5 Summary

In this analysis, we examined factors related to the fatality-counts in the 85 public mass shooting incidents (4+ victims killed in a public place) that occurred in the United States between 1995 to 2018. Our visual analytics application related to this analysis, https://mass-shooting-analytics.shinyapps.io/mass_shootings/ allows the public, policy stakeholders, and other researchers to examine and download our data, and replicate our analysis results and explore the analysis with alternate selections.

We found that the per capita rate of public mass shootings with at least four people killed has not been significantly increasing since 1995, but that the incidents became significantly more deadly. Like (Klarevas et al., 2019), we found that very high fatality incidents are becoming more frequent; for example, incidents with more than 6 fatalities per event have been increasing at close to 7% per year.

We also found that public mass shootings involving high capacity magazines are becoming more common, that such shootings have higher fatality counts, and that an increase in the use of HCM firearms followed the elimination of the federal assault weapons ban. Finally, we found that HCM pistols have increased the fatality counts as much as HCM rifles, and that prevalence of the use of high-capacity semi-automatic pistols in public mass shootings has more than doubled since the lapse of the ban, while the prevalence of the use of assault rifles has not significantly changed.

Our finding that the use of HCMs doubles the number of fatalities and more than triples the total number of people shot in public mass shootings indicates that a re-instatement of the federal ban on high-capacity magazines in pistols and rifles may reduce the fatalities in public mass shootings. Alternatively, a high tax on the sales of such magazines may also be well motivated.

## Data Availability

All data are available through the visual analytics application associated with this publication, https://mass-shooting-analytics.shinyapps.io/mass_shootings/

